# Referral protocol and teleconsultations to reduce waiting time for specialized oral medicine: a retrospective cohort study of TelessaúdeRS

**DOI:** 10.1101/2025.07.01.25330623

**Authors:** Dimitris Varvaki Rados, Carolina Dummel, Vinicius Carrard, Otávio Pereira D’Avila, Rudi Roman, Natan Katz, Erno Harzheim

**Affiliations:** TelessaúdeRS, Porto Alegre, Brazil; Federal University of Rio Grande do Sul, Porto Alegre, Rio Grande do Sul, Brazil; Federal University of Pelotas, Pelotas, Rio Grande do Sul, Brazil

**Keywords:** Referral, Oral Medicine, Primary Care

## Abstract

**Objective:** Primary health care (PHC) professionals must care for patients with oral problems while performing gatekeeping for specialized care. We explored the effect of a strategy combining referral protocols and teleconsultations on a waiting list.

**Study design:** This retrospective cohort study analyzed data from PHC to specialized oral medicine care from May/2014 to May/2016. The program started in May/2015, and referrals were divided into control and exposed groups. The median waiting time for in-person consultation and the proportion of referrals managed in PHC were analyzed for general and priority cases.

**Results:** There were 1464 referrals. The median waiting time for specialized consultation was reduced from 212 days (95%CI 196-227) to 131 days (I95%CI 95-166). For priority referrals, the control group had 37 days (95%CI 0 - 89) of median waiting time, and the exposed group had 12 days (95%CI 6 - 17).

**Conclusions:** Structured referral protocol and professional-to-professional consultations for oral disease improve referral waiting times. This benefit seems related to improving PHC efficacy.

## INTRODUCTION

Oral lesions are common and may have many causes, including benign, autoimmune, proliferative, and malignant disorders.[1] Primary Health Care (PHC) professionals (dentists and physicians) will identify most of these lesions, especially in universal healthcare systems.[2] Diagnosing and managing oral lesions are often challenging in PHC due to a lack of knowledge and experience.[3] Conversely, PHC professionals can manage some patients without referral with appropriate training and support. As such, PHC professionals have a double burden, having the duty to diagnose and treat highly prevalent oral lesions and be vigilant to suspicious lesions that need a referral to specialized care.

The balance in this gatekeeping function is central to a functional healthcare system. An excessively high amount of referrals to specialized care increases the waiting lines and delays the diagnosis of oral cancer, contributing to high morbidity and mortality.[4] Also, many referrals of patients to specialized care lack clinical data and the reasons for the consultation, besides having unclear or undefined priority levels.[5,6] Consequently, referral processes frequently result in a lack of continuity of care, delays in health assistance, dissatisfaction of users, and poorer prognosis for diseases with high morbidity and mortality. [7,8]

There are many approaches to improving PHC’s gatekeeping function. Electronic referrals, referral guidelines, and standardized format of information transferring improve the quality of the referrals.[9] Electronic systems are substituting paper-based referrals in many countries, saving costs for the health system, speeding up referrals, and reducing the risk of errors in the process.[10] Referral and triage systems are organizational structures that also play an essential role in improving access to care, bridging the gaps between levels of care, prioritizing more severe cases, and avoiding unnecessary referrals.[11,12]

In addition to the previous resources, telehealth initiatives can help improve healthcare networks, in general, and for patients with oral diseases.[13] TelessaúdeRS is a telehealth project in Brazil that aims to support PHC professionals in caring for their patients. The project has remote diagnosis, consultation, and education initiatives. A specific subproject supports the gatekeeping function for referrals from PHC to specialized care in the Rio Grande do Sul state (southern Brazil) - the RegulaSUS project (Box 1). [12]This study aims to evaluate the impact of a structured approach to PHC referrals to specialized care (the RegulaSUS strategy) for oral lesions in the waiting time for consultation and the size of the waiting list.

### Box 1.

Study context: Brazilian universal public health system and RegulaSUS strategy.

Brazil has wide dimensions and significant socioeconomic and health inequalities. Since 1988, Brazilians have had a universal public healthcare system, the Unified Health System (*Sistema Único de Saúde -* SUS), oriented and coordinated by primary care and organized as a network of services.[14] SUS still has many challenges, including limited care coordination and poor communication between primary care professionals and specialists, leading to healthcare network fragmentation.

RegulaSUS is a project designed to reduce the waiting time for consultations with specialists, prioritize care for more severe cases, and improve PHC efficacy. The Rio Grande do Sul state’s healthcare regulatory department and TelessaudeRS implemented it. The project developed and implemented referral protocols and clinical case discussions of waitlisted patients. The protocol development process follows seven steps and is described in detail in a previous publication.[12] In brief, it includes reviewing the most frequent reasons for referral, defining objective referral criteria based on clinical criteria and medical literature, and identifying high-risk patients. Additionally, professional-to-professional consultations are fostered through a toll-free telephone number. Regulatory healthcare professionals apply these criteria to waitlisted patients, and the cases with incomplete information or those that do not meet the criteria for in-person consultation are directed to teleconsulting. Through the toll-free telephone number, trained generalists and specialists offer advice for primary care professionals.

## MATERIAL AND METHODS

### Design and setting

We designed an observational, retrospective cohort study based on state electronic referral system data. This report follows the STROBE Checklist.[15] It was conducted in the Rio Grande do Sul state (Southern Brazil), where TelessaúdeRS implemented a structured referral strategy (RegulaSUS). [12]The study was approved by the local research ethics committee and followed the recommendations of the Declaration of Helsinki. Due to the nature of the data, direct patient consent was exempted.

### Participants

All referrals from the Rio Grande do Sul upstate primary care to specialized oral care in the state’s capital city from May/2014 to May/2016 were eligible. Referrals with no clinical data were excluded from this analysis.

### Variables

The electronic referral system has limited data, including patient identification, referral dates, and minimal clinical information. The regulatory decision on the referral and in-person consult date were also registered. The variables extracted from the electronic system were the age, sex (male and female) of the patient, referral and consulting date, referral reason (according to International Classification of Diseases 10th edition), and referral status in May/2016 (in line, canceled, or consult performed). Also, the decision of the regulator (cancel or authorize the consult) and priority level (two levels) attributed to the referral were extracted. Waiting time for consulting with a specialist (in general and according to priority level) was calculated by subtracting the in-person consult date and initial referral date.

The exposure (RegulaSUS strategy - see description in Box 1) was initiated in May 2015, so the referrals were divided into two periods. The control group (non-exposed) comprises referrals active from May 2014 to April 2015 (pre-implementation period); the group exposed to RegulaSUS comprises referrals active from May 2015 up to the end of follow-up (May 2016). Referrals that entered the system before the implementation but were active by May 2015 were considered exposed to RegulaSUS.

The oral medicine referral protocol is presented in Table 1, showing the three levels of referral priorities and the minimal content, which must be written in the referral letter. The original protocol also describes the presentation and associated signs and symptoms of the most common malignant, potentially malignant, benign, and proliferative lesions, which can occur in the oral cavity. As priority classification was not the norm before RegulaSUS, we retroactively classified priorities to analyze the waitlist reduction according to priority levels. This classification followed the referral protocols implemented in May 2015.

**Table 1.**
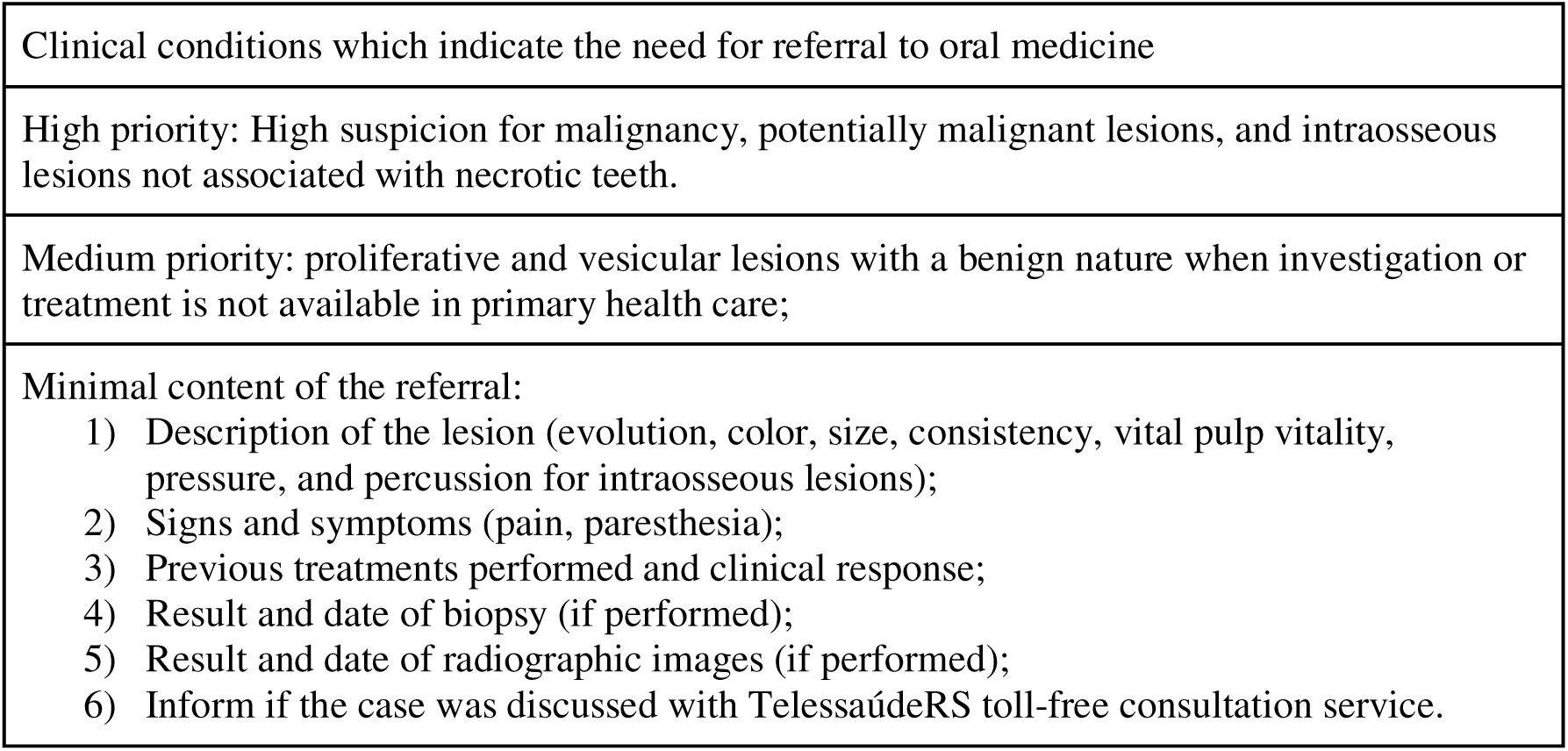
Referral protocol for oral medicine.

|  |
| --- |
| Clinical conditions which indicate the need for referral to oral medicine |
| High priority: High suspicion for malignancy, potentially malignant lesions, and intraosseous lesions not associated with necrotic teeth. |
| Medium priority: proliferative and vesicular lesions with a benign nature when investigation or treatment is not available in primary health care; |
| Minimal content of the referral:<br>1) Description of the lesion (evolution, color, size, consistency, vital pulp vitality, pressure, and percussion for intraosseous lesions);<br>2) Signs and symptoms (pain, paresthesia);<br>3) Previous treatments performed and clinical response;<br>4) Result and date of biopsy (if performed);<br>5) Result and date of radiographic images (if performed);<br>6) Inform if the case was discussed with TelessaúdeRS toll-free consultation service. |

### Sample size and data sources

All referrals to specialized oral medicine consults in the Rio Grande do Sul state from May 2014 to May 2016 were eligible. Anonymous data from the referrals was extracted from the state’s electronic system.

### Data analysis

The descriptive analysis was conducted to present patients’ characteristics and referrals. We explored RegulaSUS exposure (before and after implementation) on waiting time with survival analysis. Time for consultation was assessed with Kaplan-Meyer analysis for general and priority referrals. To further explore the performance of RegulaSUS, we also analyzed patients with malignant and premalignant disease codes that the referring professional provided. Data were analyzed using PASW 18.0 for Windows (SPSS, Inc., Chicago, IL).

## RESULTS

In total, 1464 referrals were included in the study; 586 were considered in the control group and 878 in the exposed group. By May 2014, 375 referrals were waiting for specialized consultation, and 495 new referrals were included in the system between May 2014 and April 2015. At the beginning of RegulaSUS strategy for oral medicine (May 2015), there were 288 active referrals in line, and 594 new referrals were performed up to May 2016. Figure 1 shows the study flow diagram. No referral was excluded from the primary analysis; however, 51 referrals were excluded from the analysis stratified by priority due to insufficient clinical data. Regarding patients’ characteristics, the mean age (52.3 ± 18.6 for the before group and 51.8 ± 17.5 for the after group) and proportion of females (61.5% in the before group and 57.9% in the after group) were similar. The most common referral reasons, according to the International Classification of Diseases informed by the PHC professional, were lip (18%), tongue (6%), and gingival (6%) lesions. Aftous lesions were reported in 4% of referrals, and cancer-related referrals comprised 3% of the sample. The distribution was similar between the groups. In the priority evaluation, the control group had fewer high-risk referrals (3.6% vs. 8.7%).

**Figure 1.**
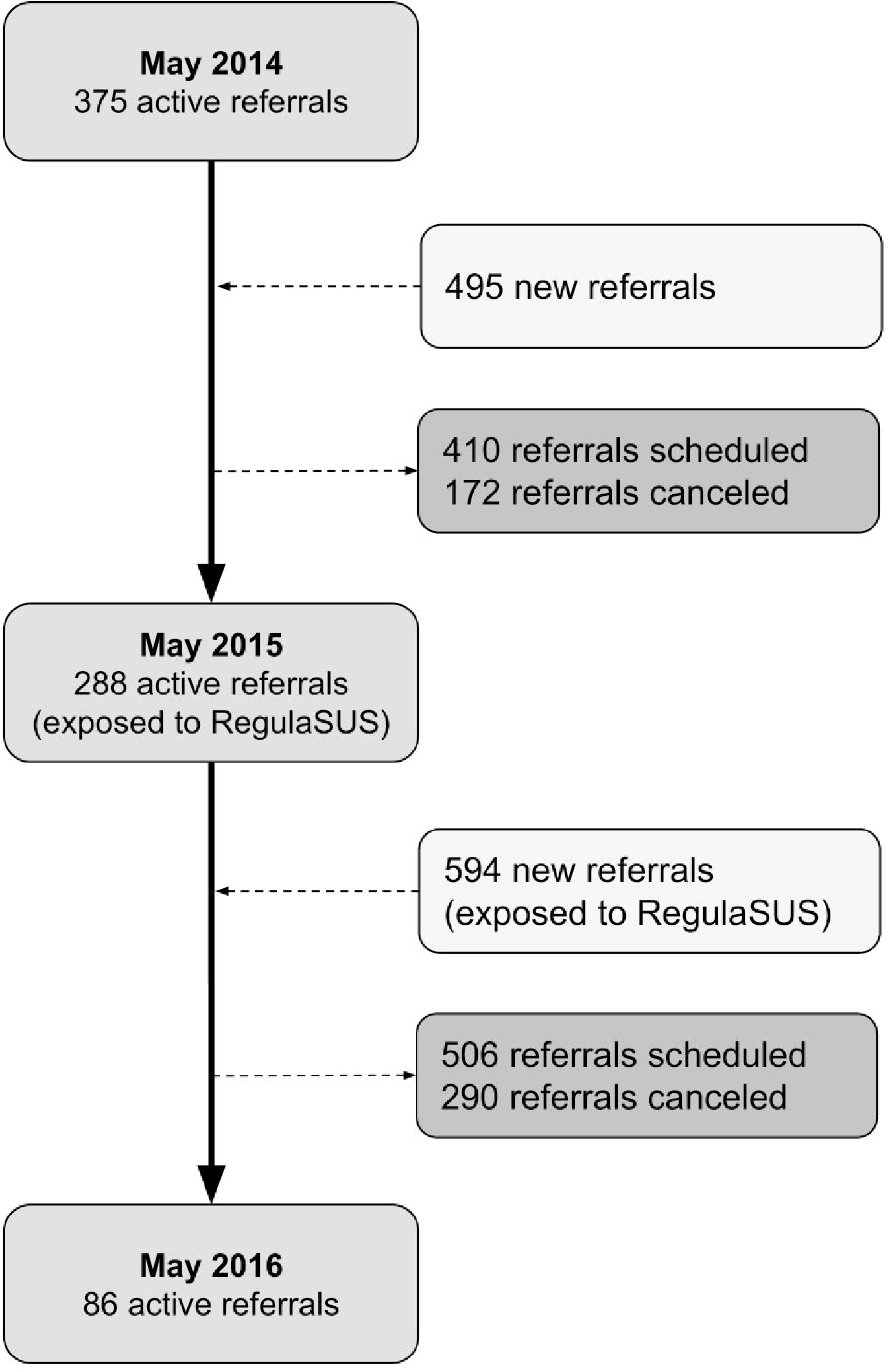
Study flow diagram.

Figure 1 shows that 410 patients were scheduled for in-person consults in the control group and 506 in the exposed group. In the overall analysis, the median time for in-person consultations was shorter for the referrals exposed to RegulaSUS (212 [95%CI 196 - 227] vs. 131 days [95%CI 95 - 166], p< 0.001; before and after groups, respectively). Figure 2 shows the proportion of patients being (one-minus survival) scheduled over time. When the dataset was restricted to priority referrals, there were 21 consults before and 71 after RegulaSUS began. Regarding the median time for consultations, the control group had 37 days (95%CI 0 - 89), and the exposed group had 12 days (95%CI 6 - 17) (p=0.01). In the analysis of suspected malignant and premalignant referrals, similar results were identified: before RegulaSUS, this subgroup had a median waiting time of 222 days (95%CI 151 - 292), and it was reduced to 24 days (95%CI 13 - 34) after implementation (p<0.001).

**Figure 2.**
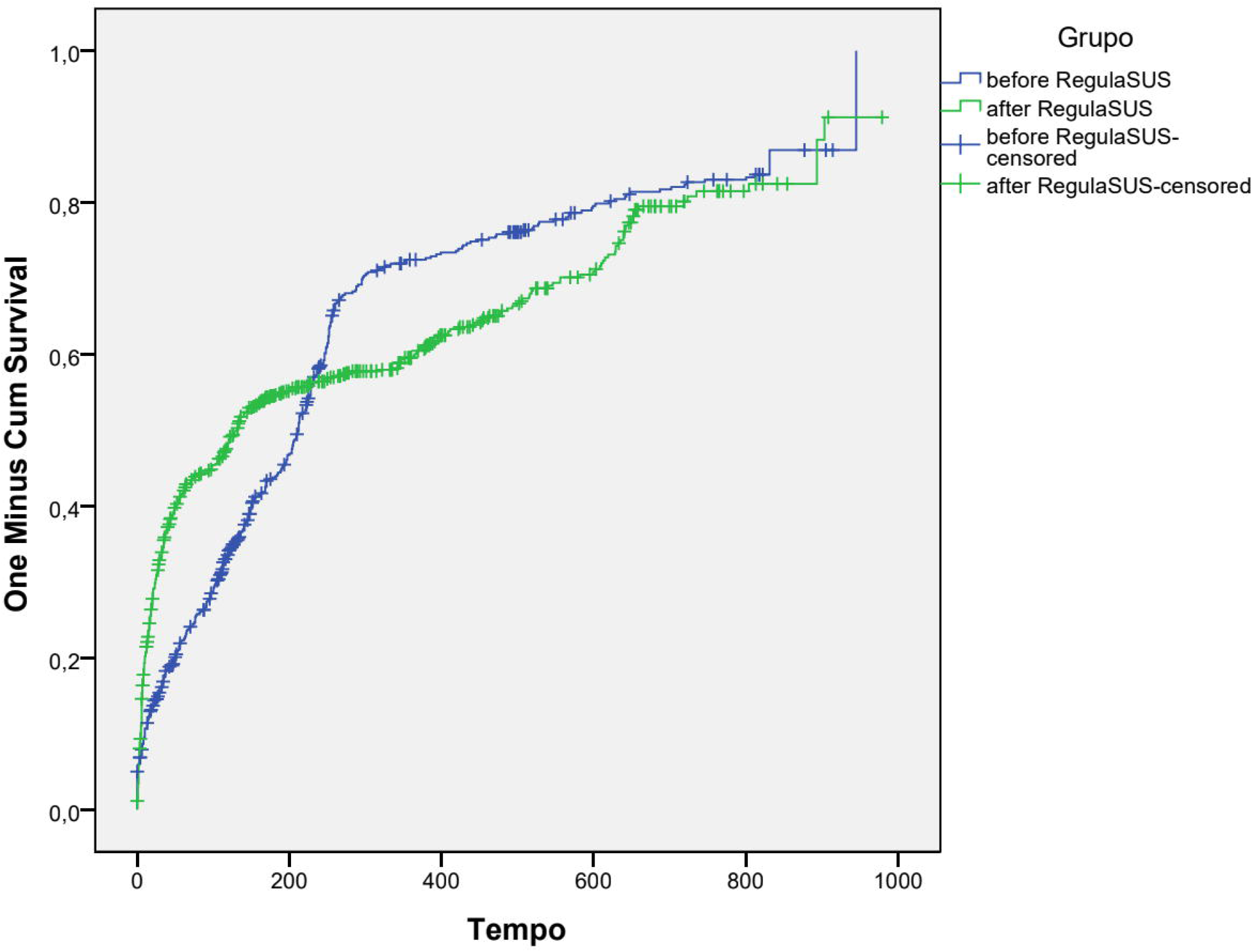
Kaplan-Meier one minus survival curve for in-person consults scheduling.

## DISCUSSION

The present study analyzed the impact of a structured approach to gatekeeping referrals from PHC (RegulaSUS) on the waiting time for specialized oral medicine consultations. Our results show that this approach reduces waiting time by almost 100 days. This benefit seems even more significant in high-priority or suspected malignancy cases, where the waiting time is reduced manyfold.

The benefits identified in this study favor both patients and the healthcare system. By reducing the waiting time, patients needing oral medicine in-person evaluation reach specialists in adequate time. This improvement may have relevant additional benefits. By improving primary care efficacy and avoiding unnecessary in-person visits, higher patient satisfaction may be achieved, fewer travels will be needed, and reduced overcrowding of specialized care (high-cost resources) can be achieved. Additionally, high-risk cases (elevated suspicion or confirmed cancer) benefit the most. This benefit may improve prognosis, as diagnosis and treatment delays for oral cancer directly impact survival rates.[16]

It is known that protocols are helpful in the reduction of erroneous or lacking information and referrals and help prioritize patients with more urgent cases, improving overall healthcare system quality.[17] RegulaSUS combines protocols with an approach to waitlisted patients, offering professional-to-professional consultations and improving PHC to care for patients without in-person referrals. This may explain the benefits of the initiative, as other experiences with consultations (combined or not with referrals) show variable benefits. Consultations with pediatricians avoided in-person visits and hospital care. [18] A Canadian experience shows that written e-consultations avoided almost 50% of referrals across 16 specialties. [19] Similar to our results and clinical context, a systematic review showed that referral protocols for prioritization improve waiting times for surgical specialties.[20] Finally, a systematic review of asynchronous e-consultations showed worldwide experience with this kind of service; however, most studies reported avoidance of in-person visits. [21]

The design of our study has intrinsic limitations. Due to the retrospective and observation nature of the available data, the causal relation between exposure to the RegulaSUS strategy and the reduced waitlist has yet to be confirmed. Despite this, the relationship between new referrals and consultations was stable. Therefore, the benefits can not be attributed to increased oral medicine consults. Other data from the same strategy in other referral lists reinforce our results.[12,22] Although long waiting lists occur in various countries,[23] another significant limitation is that this system was developed and implemented in a specific context in a state of a middle-income country with a healthcare system with universal coverage. Previous data show that avoidance of in-person visits varies widely (7.8% to 78%). As such, the results presented here may not be reproducible in other health and economic contexts.

Some cardinal aspects must be included when developing a similar strategy in other contexts to reach similar results. First, referral criteria must be defined to address the most common and high-risk clinical problems, as shown in Table 1 and described elsewhere. [12] Second, these criteria must be applied for waitlisted referrals, and those eligible must be authorized for in-patient care. Third, patients not eligible for specialized in-person care (or with unclear data) must be directed to remote consultations, irrespective of the modality (telephone, asynchronous, text-based).

Unfortunately, we could not evaluate direct patient outcomes related to the disease, satisfaction, or quality of care provided. We may hypothesize that decreasing waiting times will provide additional benefits for future referrals (earlier diagnosis and better treatment results); however, we had no access to clinical data to confirm this. This limitation must be acknowledged when considering the generalization of our findings, as our study does not answer additional relevant effects for patients (besides waiting time).

We also acknowledge that not every canceled referral can be attributable to improved PHC efficacy. There are many other reasons for excluding a referral from the waiting list, such as death, patient unreachable, or self-limited disease. Despite this, it seems unlikely that our intervention affected these events. In other words, even considering the other reasons for being canceled, most of the patients excluded from the referral line by the RegulaSUS strategy are probably attributable to being cared for in the PHC.

In summary, RegulaSUS helped to reduce the waiting time for specialized consultation, especially for high-risk patients. It also assisted PHC professionals in managing patients not fulfilling the protocol criteria. Our findings support referral protocols aligned to consultations to improve PHC and the referral process.

## Data Availability

The datasets used and/or analyzed during the current study are available from the corresponding author on reasonable request and clear analysis plan.

## Authors’ Contributions

DVR.: conceptualization, data curation and analysis, writing - original draft and editing.

VC: conceptualization, writing - original draft.

OPD: conceptualization, writing - original draft.

RR: conceptualization, data curation, writing - review.

NK: conceptualization, data curation, writing - review.

EH: initial conceptualization, data curation, writing - review.

All authors approved the final version of the article as submitted and agree to be accountable for all aspects of the work.

## Acknowledgments

The authors are grateful to the Rio Grande do Sul state health authority for the support and financing of the whole RegulaSUS strategy.

## REFERENCES

1. Zahid E, Bhatti O, Zahid MA, Stubbs M. Overview of common oral lesions. Malaysian Family Physician : the Official Journal of the Academy of Family Physicians of Malaysia. 2022;17: 9.

2. Grafton-Clarke C, Chen KW, Wilcock J. Diagnosis and referral delays in primary care for oral squamous cell cancer: a systematic review. Br J Gen Pract. 2019;69: e112.

3. Strey JR, Roxo-Gonçalves M, Guzenski BD, Martins MAT, Romanini J, de Figueiredo MAZ, et al. Oral Medicine Experience and Attitudes Toward Oral Cancer: An Evaluation of Dentists Working in Primary Health Care. J Cancer Educ. 2022;37. doi:10.1007/s13187-021-01999-z

4. Torres-Pereira CC, Angelim-Dias A, Melo NS, Lemos CA Jr, Oliveira EMF de. Abordagem do câncer da boca: uma estratégia para os níveis primário e secundário de atenção em saúde. Cad Saúde Pública. 2012;28: s30–s39.

5. Senitan M, Alhaiti AH, Gillespie J, Alotaibi BF, Lenon GB. The Referral System between Primary and Secondary Health Care in Saudi Arabia for Patients with Type 2 Diabetes: A Systematic Review. J Diabetes Res. 2017;2017: 4183604.

6. Esan O, Oladele O. Referral letters to the psychiatrist in Nigeria: is communication adequate? Afr Health Sci. 2016;16: 1023–1026.

7. Hartveit M, Vanhaecht K, Thorsen O, Biringer E, Haug K, Aslaksen A. Quality indicators for the referral process from primary to specialised mental health care: an explorative study in accordance with the RAND appropriateness method. BMC Health Serv Res. 2017;17: 4.

8. 8. Almeida PF de, Santos AM dos. Primary Health Care: care coordinator in regionalized networks? Rev Saúde Pública. 2016;50: 80.

9. Maghsoud-Lou E, Christie S, Abidi SR, Abidi SSR. Protocol-Driven Decision Support within e-Referral Systems to Streamline Patient Consultation, Triaging and Referrals from Primary Care to Specialist Clinics. J Med Syst. 2017;41: 139.

10. Heimly V. Electronic referrals in healthcare: a review. Stud Health Technol Inform. 2009;150: 327–331.

11. Kapoor R, Avendaño L, Sandoval MA, Cruz AT, Sampayo EM, Soto MA, et al. Initiating a Standardized Regional Referral and Counter-Referral System in Guatemala: A Mixed-Methods Study. Glob Pediatr Health. 2017;4: 2333794X17719205.

12. Katz N, Roman R, Rados DV, Oliveira EB de, Schmitz CAA, Gonçalves MR, et al. Access and regulation of specialized care in Rio Grande do Sul: the RegulaSUS strategy of TelessaúdeRS- UFRGS. Ciênc saúde coletiva. 2020;25: 1389–1400.

13. Tagliaferri S, Esposito FG, Ippolito A, Mereghini F, Magenes G, Martinelli P, et al. Telemedicine to Improve Access to Specialist Care in Fetal Heart Rate Monitoring: Analysis of 17 Years of TOCOMAT Network Clinical Activity. Telemed J E Health. 2017;23: 226–232.

14. Macinko J, Harris MJ. Brazil’s family health strategy--delivering community-based primary care in a universal health system. N Engl J Med. 2015;372: 2177–2181.

15. von Elm E, Altman DG, Egger M, Pocock SJ, Gøtzsche PC, Vandenbroucke JP, et al. The Strengthening the Reporting of Observational Studies in Epidemiology (STROBE) statement: guidelines for reporting observational studies. J Clin Epidemiol. 2008;61: 344–349.

16. Chiou S-J, Lin W, Hsieh C-J. Assessment of duration until initial treatment and its determining factors among newly diagnosed oral cancer patients: A population-based retrospective cohort study. Medicine . 2016;95: e5632.

17. Olivotto IA, Gomi A, Bancej C, Brisson J, Tonita J, Kan L, et al. Influence of delay to diagnosis on prognostic indicators of screen-detected breast carcinoma. Cancer. 2002;94: 2143–2150.

18. Wegner SE, Humble CG, Feaganes J, Stiles AD. Estimated savings from paid telephone consultations between subspecialists and primary care physicians. Pediatrics. 2008;122. doi:10.1542/peds.2008-0432

19. Keely E, Liddy C, Afkham A. Utilization, benefits, and impact of an e-consultation service across diverse specialties and primary care providers. Telemedicine journal and e-health : the official journal of the American Telemedicine Association. 2013;19. doi:10.1089/tmj.2013.0007

20. Rathnayake D, Clarke M. The effectiveness of different patient referral systems to shorten waiting times for elective surgeries: systematic review. BMC Health Services Research. 2021;21: 1–13.

21. Liddy C, Moroz I, Mihan A, Nawar N, Keely E. A Systematic Review of Asynchronous, Provider-to-Provider, Electronic Consultation Services to Improve Access to Specialty Care Available Worldwide. Telemedicine journal and e-health : the official journal of the American Telemedicine Association. 2019;25. doi:10.1089/tmj.2018.0005

22. Pfeil JN, Rados DV, Roman R, Katz N, Nunes LN, Vigo Á, et al. A telemedicine strategy to reduce waiting lists and time to specialist care: A retrospective cohort study. J Telemed Telecare. 2023;29: 10–17.

23. Amigoni F, Lega F, Maggioni E. Insights into how universal, tax-funded, single payer health systems manage their waiting lists: A review of the literature. Health Serv Manage Res. 2023 [cited 19 Aug 2023]. doi:10.1177/09514848231186773

